# RSV concentration in wastewater and trends among incident hospitalizations in children and older adults

**DOI:** 10.64898/2026.07.21.26358587

**Authors:** Dustin T. Hill, Jennifer Laplante, Sylvia Byun, Mohammed Alazawi, Donna Gowie, Sonali Biswas, Latavia Hill, Yifan Zhu, Monica R. Foote, Haley Kappus-Kron, Milagros Neyra Blatz, Ian Bradley, Yinyin Ye, Kirsten St. George, David A. Larsen

## Abstract

Wastewater-based epidemiology (WBE) has the potential to fill gaps in clinical surveillance for respiratory syncytial virus (RSV) burden. Previous studies have shown RSV to be detectable in wastewater, but few have linked detections to clinical data because, in many locations, RSV is not a reportable disease. Further, studies that have linked RSV in wastewater to clinical data have not distinguished between pediatric and adult infections, as is common in studies of RSV. Using 2,662 influent wastewater samples collected from twenty-four treatment plants in four New York counties, we measured the RSV concentration in wastewater and compared levels detected to RSV hospitalizations from September 2022 to July 2024. RSV concentrations in wastewater correlated well with RSV hospitalizations (ρ between 0.52 and 0.85, *P* <0.001). RSV concentrations in wastewater lagged hospitalizations for under 10-year-olds by an average of three weeks but were a leading indicator for hospitalizations in patients over 50 by up to two weeks. In predictive models, wastewater explained 88 percent of the variance in RSV hospital admissions among people over 50. Wastewater-based epidemiology of RSV is therefore a reliable surveillance method and could provide early warning for increases in RSV hospitalizations among older populations.

## 1 Introduction

Respiratory syncytial virus (RSV) is a seasonal respiratory virus that afflicts millions of individuals annually^1^. RSV has two major subgroups designated as RSV A and RSV B^2^. RSV infections are usually seasonal with a rise in cases during autumn months in temperate, northern hemisphere climates, peaking during the winter between December and February followed by a decline in new infections in the spring^2^. Most children experience RSV at least once by the age of two^3^, usually with symptomatic presentation^4^, and between 15-50% of infected children develop lower respiratory tract infections such as bronchitis, pneumonia, or croup, often requiring hospitalization^5^. Infection with RSV can lead to severe manifestations requiring hospitalization, which is most common among children and older adults^6,7^.

Older children and adults (aged >50 years) may be reinfected with RSV throughout their lives but this typically results in far milder symptoms, with the infection remaining in the upper respiratory tract^8^. Adults over the age of 50, particularly those with underlying medical conditions may be at elevated risk of reinfection and the potential for more serious RSV disease and possibly hospitalization^9^. Up to half of all RSV hospitalizations in a given season are among older adults^10^ and RSV can cause up to ten percent of hospitalized pneumonia cases in older adults^11^

It is not definitively known what drives RSV transmission, although annual seasonal patterns have been linked to climate and environmental factors^12^. Whether older or younger adults drive RSV transmission is not immediately clear, however, research on the mixing of populations has found that contact rates between infants and the elderly is generally the lowest among different age groups^13^. Studies that investigated younger versus older RSV infections have found that hospitalizations tend to rise first among children before rising in older adults^14^.

RSV seasons can be monitored through clinical reports and with wastewater surveillance^15,16^, however, wastewater surveillance is a relatively new method for following the disease and has only seen wider use since 2020^17^. Most wastewater-based epidemiological (WBE) studies of RSV have not investigated the relationships to clinical data^16,18^ with some indicating that clinical data were not available since RSV was not a reportable disease in the areas studied and the virus presented clinical symptoms similar to other respiratory pathogens^19^. Wastewater has the advantage of providing community-level data on disease levels with a single sample that could be a good indicator of disease transmission when clinical surveillance is less reliable^20^. Previous studies that have tested RSV in wastewater have found that detection levels are highest during the winter months, which would match the typical respiratory season for clinical cases^21^. Wastewater testing of RSV has also shown that the subtypes most prevalent in clinical cases match the subtypes detected in the wastewater^22^.

In this study, we analyze RSV wastewater concentration data collected in New York State (NYS) between 2022 and 2024 from four counties (Erie, Jefferson, Onondaga, and Westchester) and compare those data with reported clinical cases and hospitalizations. We further explore if the relationship between RSV in wastewater and clinical data differs across different subgroups of susceptible people by assessing infections in children and older adults separately.

## 2 Methods

### 2.1 Wastewater data

Wastewater samples were collected from four counties (Erie, Jefferson, Onondaga, and Westchester) in NYS across 27 WWTP influent points within the New York State Wastewater Surveillance Network^23^ from September 2022 to July 2024. Samples were collected once or twice a week and were shipped on ice to one of two labs (Quadrant Biosciences and University at Buffalo, SUNY or UB) for nucleic acid extraction, extracts were then shipped to the Virology Lab at Wadsworth Center (hereafter Wadsworth) in Albany, New York for digital PCR (dPCR) analysis. Samples were divided up by lab where Quadrant Biosciences used ultracentrifugation^24^ to concentrate samples collected from Jefferson, Onondaga, and Westchester counties. Briefly, 20 mL wastewater sample and 12 mL sucrose cushion were added to an ultracentrifuge cube. The mix was then ultracentrifuged at 150,000 x *g* at 4°C for 45 minutes. The pellet was resuspended in 200 µL 1X PBS, and nucleic acid extraction was performed using the AllPrep® PowerViral® DNA/RNA Kit (Qiagen, Germany) with a final elution volume of 50 µL. UB used Nanotrap® Microbiome Particles (Ceres Nanosciences, USA) to concentrate virus particles from wastewater samples collected from Erie County. Briefly, 9.75 mL wastewater sample was mixed with 50 µL Nanotrap® Enhancement Reagent 1 and 75 µL Nanotrap® Microbiome A Particles. The virus particles were eluted in 500 µL of MagMAX™ Microbiome Lysis Solution. Then, 400 µL of the lysates were mixed with 530 µL of MagMAX™ Binding Solution, 10 µL of MagMAX™ Proteinase K, and 20 µL of MagMAX™ DNA/RNA Binding Beads. The nucleic acids were washed in MagMAX™ Wash Buffer and then 80% ethanol, finally eluted in 50 µL of MagMAX™ Elution Buffer. Both concentration and nucleic acid extraction were performed using the KingFisher™ Apex Purification System (ThermoFisher Scientific, USA). More details of the concentration and nucleic acid extraction methods can be found in previously published literature^25^.

### 2.2 RSV quantification in wastewater

Wadsworth developed and validated a multiplexed dRT-PCR assay for influenza A virus and respiratory syncytial virus (RSV) detection and quantitation in wastewater. The assay targets the highly conserved regions of the matrix gene of influenza A virus and the matrix gene of RSV (types A and B). RSV primer and probe sequences were derived from Fry et al^26^, with modifications to the reverse primer (RSV-pan-R 5’TCTYTTTCTAGRACATTGTAYTGRACAG3’) to enhance the detection of currently circulating RSV viruses. The dRT-PCR reactions of the multiplexed assay were optimized using the QIAcuity OneStep Advanced Probe Kit (QIAGEN). The final 50µL volume includes 12.5µL of 4X QIAcuity One-Step MM, 0.5µL of 100X Qiagen One Step RT-PCR Mix, 5µl of total nucleic acid, a mix containing four primers at 900nM each and one FAM probe at 250nM, targeting influenza A, along with two primers at 900nM each and one HEX probe at 250nM targeting RSV. Each probe is synthesized with a ZEN internal quencher set between the 9^th^ and 10^th^ nucleotides, and Iowa Black quencher (Integrated DNA Technologies, Inc). The QIAcuity (QIAGEN) cycling conditions include 30 minutes at 50°C, 2 minutes at 95°C, and 45 cycles of 15 seconds at 95°C and 30 seconds at 55°C. Assay specificity was evaluated with a blinded panel of twenty-two respiratory viruses, including various strains of influenza A, RSV, SARS-CoV-2, adenovirus, enterovirus, parainfluenza virus, rhinovirus, influenza B, varicella zoster virus, and cytomegalovirus. No cross reactivity was observed. Additionally, the assay’s limit of detection was assessed using quantitated influenza A/H1pdm09 and A/H3 viruses and RSV A and B viruses. Viruses were tested individually and in varying known mixed concentrations. The assay reliably detects single and mixed influenza A and RSV, to concentrations as low as 1 and 10 gene copies/µL of reaction respectively. The 26k QIAcuity nanoplate was used for all dRT-PCR validation and wastewater testing. Viral gene copies were quantified by the QIAcuity instrument. All results were converted back to the concentration in wastewater samples.

### 2.3 Case data

RSV case data were collected from the National Respiratory and Enteric Virus Surveillance System (NREVSS). NREVSS is a clinical laboratory-based network with participating healthcare centers around the United States. Data were collected between September 13, 2022, and July 10, 2024. In this study only Jefferson and Onondaga counties overlapped with the NREVSS network. Data were summed to the county level from participating facilities. RSV A, RSV B, and RSV unspecified test results were combined to create one variable for total reported RSV cases for each county. Incidence was calculated using county population to convert to positive tests per 100,000 population, and population of each age group in the county (e.g., children under 10).

### 2.4 Hospitalization data

RSV hospitalization data were obtained from the Statewide Planning and Research Cooperative System (SPARCS). The hospitalization dataset from SPARCS contained longitude and latitude coordinates of the patient addresses. We refined the dataset by removing records with missing latitude or longitude information and those that extended beyond the borders of New York State (i.e., latitude beyond 40 and longitude beyond −80) were excluded from the analysis. Further exclusions were made on cases that were not spatially linked to mapped sewersheds using the Simple Feature (sf) package in R, excluding those that fell outside the sewershed areas. These were then assigned to WWTPs via the sewershed boundary associated with each WWTP as has been previously described for COVID-19^25,27^. Hospital admissions on the same day were summed to the sewershed collection area of their residence providing an estimate for total admissions on any given day. Hospital admissions were also summed across the county for the pilot counties and included all residents even if they did not reside within a sewershed.

To estimate RSV hospitalizations, all ICD 10 codes were combined for RSV Pneumonia, RSV Acute Bronchiolitis, and RSV since the same patients were diagnosed with multiple codes on the same date. See supplemental materials for specific ICD 10 codes for RSV (Table S4).

We also obtained counts of RSV hospital admissions for two sub-populations. Since RSV hospitalizations are most common in young children and older adults, we obtained a dataset for admissions of under 10-year-olds and another dataset for over 50-year-olds. This allowed for comparing the wastewater data to all hospitalizations, those under 10, and those over 50. Under 10-year-olds was selected as a category because the majority of childhood RSV hospitalizations occur for under 2-year-olds^28^, but we wanted to include other children that might have infection in later years because of lack of exposure during the COVID-19 pandemic^29,30^. Population data were used to further compute incidence of hospital admissions per 100,000 for sewersheds and counties. Age-adjusted incidence for under 10-year-olds and under-50-year-olds was computed using countywide estimates for these age groups. Countywide estimates were obtained from “tidycensus”^31^ and the data retrieval was done with the assistance of Claude AI.

### 2.5 Statistical analysis

Data were transformed into a weekly time series since most WWTPs sampled once per week. For weekly aggregation of sites that sampled twice weekly, wastewater concentration was averaged across samples. To obtain county level estimates, data were population weighted by week to the county level using the population of the sewershed served by the WWTP.

For case data, days with no reported cases were substituted with zero and then a weekly sum was calculated. County-level estimates were obtained by summing all the RSV cases in the county from NREVSS reporting sites.

For hospitalization data, days with no reported hospitalizations were substituted with zero and then a weekly sum was calculated. County-level estimates were obtained by summing all the reported hospital admissions in the county. We conducted analyses of data reported during the entire year (all weeks) as well as during respiratory season (weeks 40 to 20)..

Statistical correlation between weekly average wastewater concentration and weekly case incidence and hospitalization incidence for RSV were estimated using Spearman’s rank correlation. Spearman was selected because of nonlinearity in the data even after a log+1 transformation.

Where results were substantially different, we report results for both in the main text. Where results were not substantially different, we report results in the supplementary materials Sensitivity, specificity, positive predictive value (PPV) and negative predictive value (NPV) were estimated for RSV wastewater concentration and related RSV hospitalizations. We defined a true positive as a detection of RSV in wastewater with a corresponding hospital admission or positive PCR case in the same week.

We also assessed whether the wastewater signal preceded the reporting of a positive test or hospital admission by testing if lagged wastewater data had higher correlation with observed case/hospitalization numbers from the weeks leading the case/hospitalization report or if the signal lagged the clinical data. We tested this for up to eight weeks before and after the week of the reported clinical data. We conducted a second analysis with the clinical data splitting RSV hospital admissions into the two groups with the highest burden, children under 10 and adults over 50 years old. Case data were not split because age was not available from NREVSS data.

We also tested the predictive ability of weekly RSV wastewater concentration on weekly RSV hospitalizations for over 50-year-olds in time lagged models. We fit three sets of models for lags of zero to two weeks using a generalized linear mixed model (GLMM) with a gaussian error structure. We included county as a random effect and used an autoregressive error structure to account for the repeated measures of the weekly data. For each of the four lagged models we fit versions with the two predictors lagged wastewater concentration and lagged RSV hospital admissions for under 10-year-olds. We also fit versions with wastewater concentration only and RSV hospital admissions only as predictors. Models were fit in R package “glmmTMB”^32^ and all analyses were conducted in R version 4.5.1^33^. Besides using Claude AI to assist with the data retrieval from “tidycensus”, Claude AI was used to help write the R code for table formatting. Generative AI was not used for data analysis.

## 3 Results

### 3.1 RSV infection rates and wastewater detection rates

In total, 2,662 wastewater samples were collected during two respiratory seasons, from twenty-four sites across four counties, and tested for RSV (Figure 1). There was an average detection rate between 17.8% and 41.1% across all sites throughout the study period (Table 1). Erie County had the highest detection rate with one site reporting positive results in 59.1% of samples (Figure 1, Table 1), while Onondaga County had the lowest detection rate of 14.3% (Figure 1, Table 1). Erie County samples also demonstrated the highest virus concentrations ranging from 0 to 165.23 copies per mL from the magnetic bead extracted samples, while the three other counties using the ultracentrifugation processing method exhibited levels ranging between 0 and 85.35 copies per mL Table 1). During the respiratory season, detection rates were higher: in Erie County they averaged 56.58% as opposed to 41.12% across the entire year, in Jefferson County 22.37% compared to 17.80%, Onondaga County 36.53% compared to 30.44%, Westchester County 44.37% as opposed to 38.14% during all weeks of the year (Table S1).

**Figure 1:**
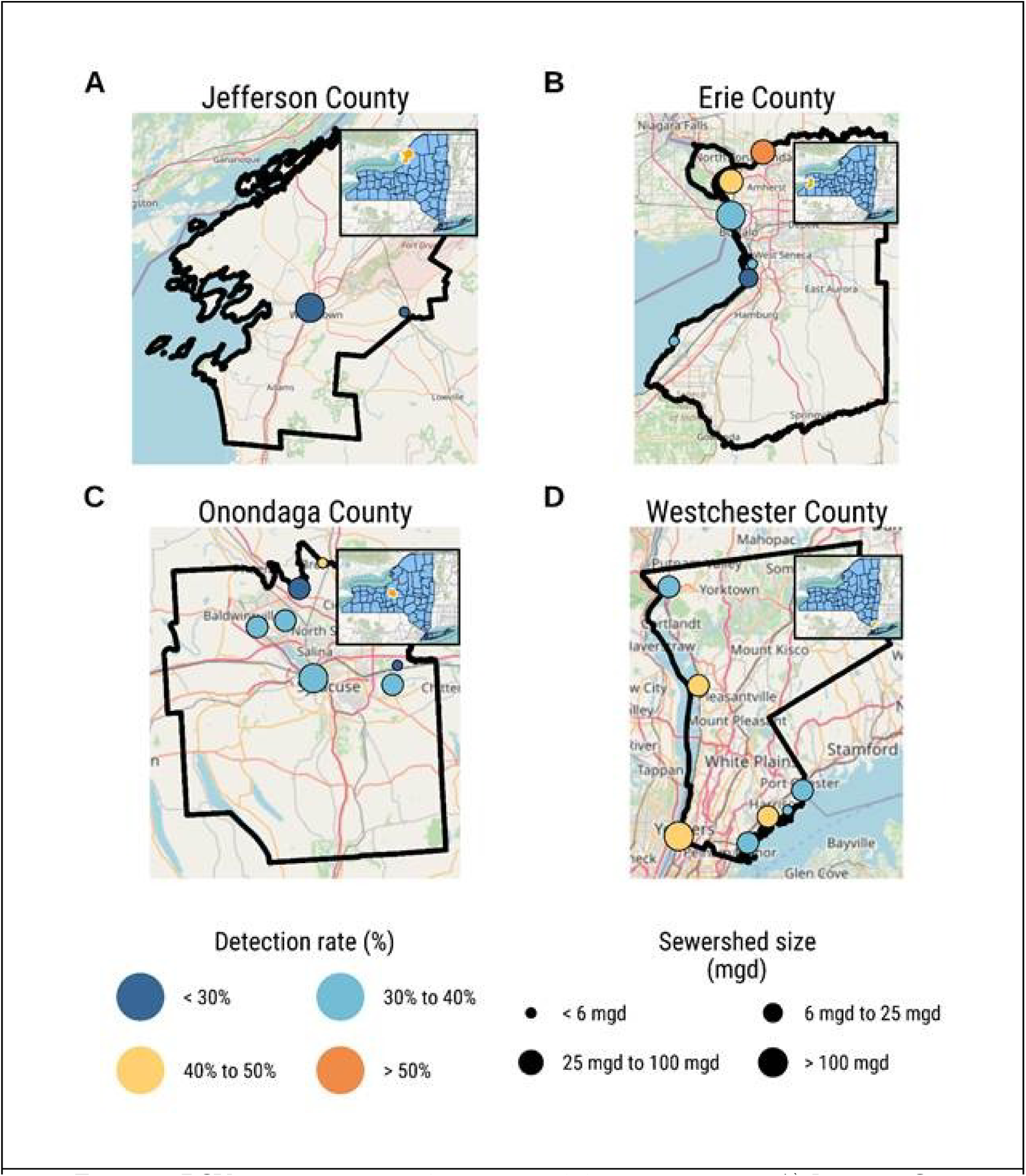
RSV wastewater sampling sites and detection rates for A) Jefferson County, B) Erie County, C) Onondaga County, and D) Westchester County.

**Table 1:**
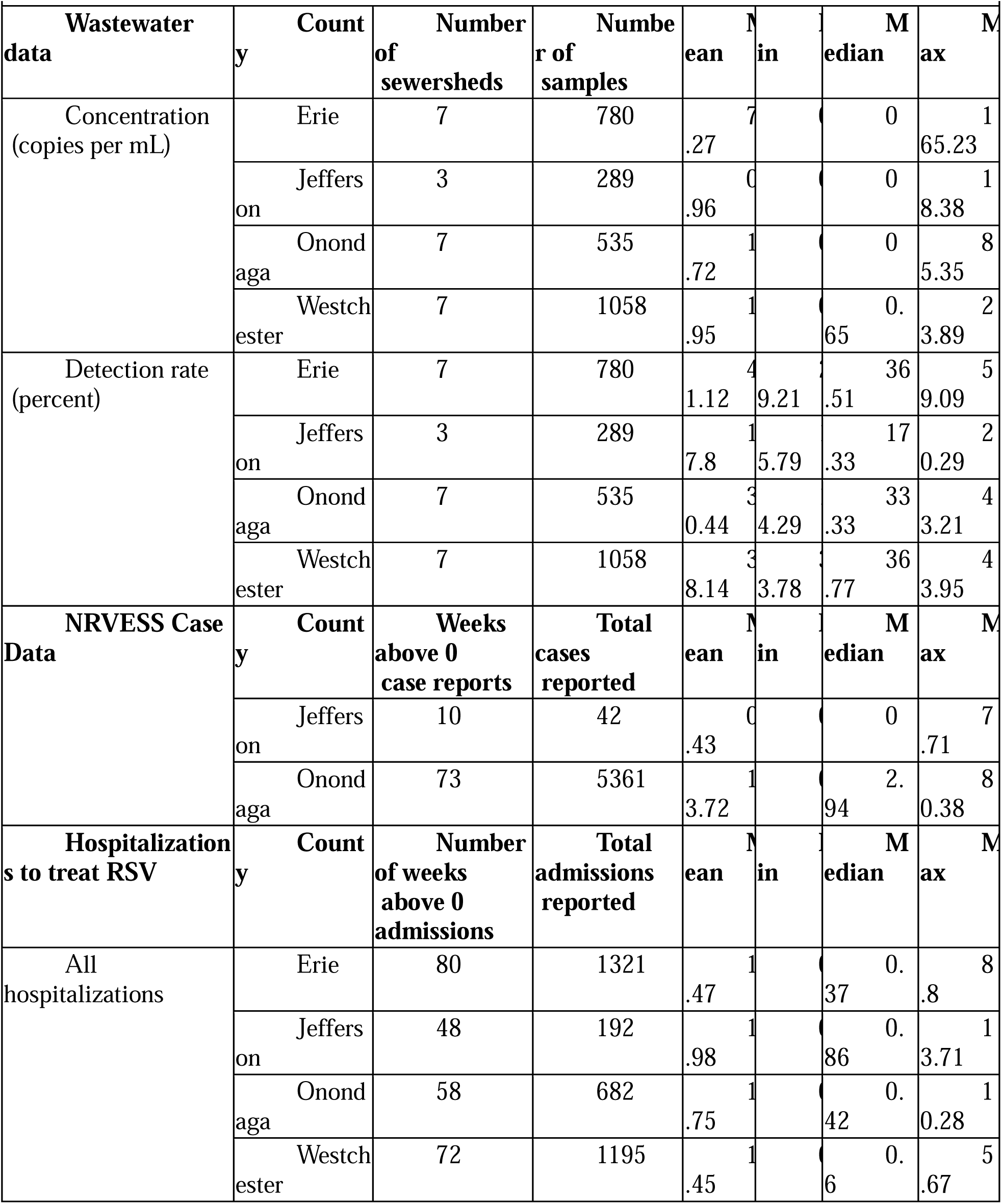
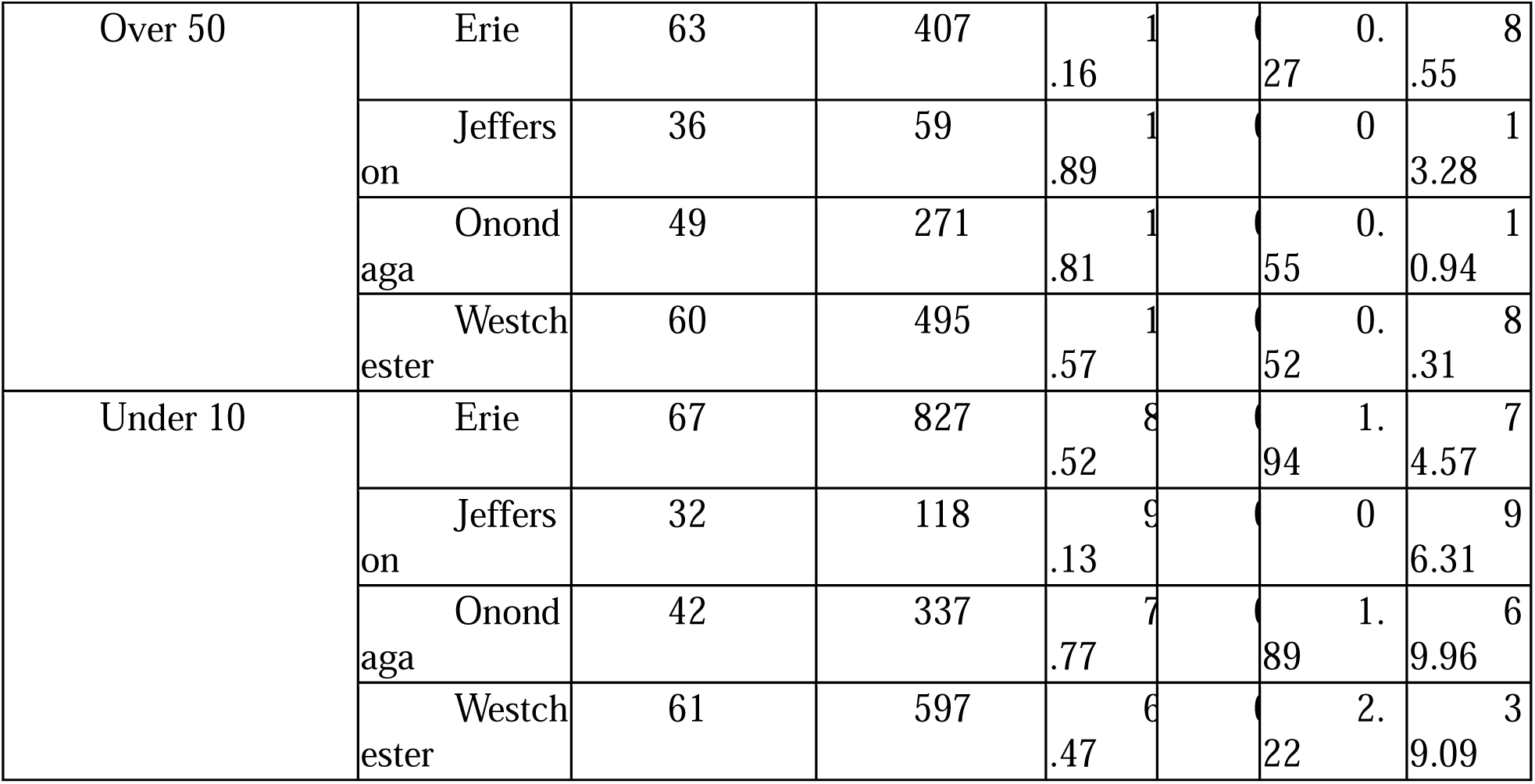
Descriptive statistics. Values are for RSV wastewater concentration data from each sample, reported positive tests for RSV in people (cases), and reported hospital admissions for treating RSV.

RSV positive clinical cases were reported in Onondaga and Jefferson Counties, while Erie and Westchester Counties did not participate in NREVSS surveillance (Table 1). Onondaga County reported the most RSV cases over the two respiratory seasons with 5,361 reports and an average incidence of 13.72 per 100,000 population (Table 1). In contrast, Jefferson County only reported 42 cases and an average incidence of 0.43 cases per 100,000 with testing only occurring in only one town in the county. RSV hospital admissions were reported in all counties with the most admissions reported in Erie County at 1,321 with an average incidence of 1.47 admissions per 100,000 during both seasons (Table 1). Jefferson County had the least number of hospitalizations with 192 reported over the two seasons and an average incidence of 1.98 per 100,000 population during both seasons (Table 1).

### 3.2 Detection trends of RSV in wastewater

RSV concentration in wastewater in each county showed the same general trend peaking at similar times during each respiratory season: in December 2022 in the 2022-2023 season and in late December 2023 in the 2023-2024 season (Figure 2). RSV cases and hospital admissions peaked at similar times within the counties, but the clinical reports appeared to peak before the peak wastewater signal (Figure 2). This was evident in each county for both RSV case data and hospitalizations with some variation between seasons (Figure 2).

**Figure 2:**
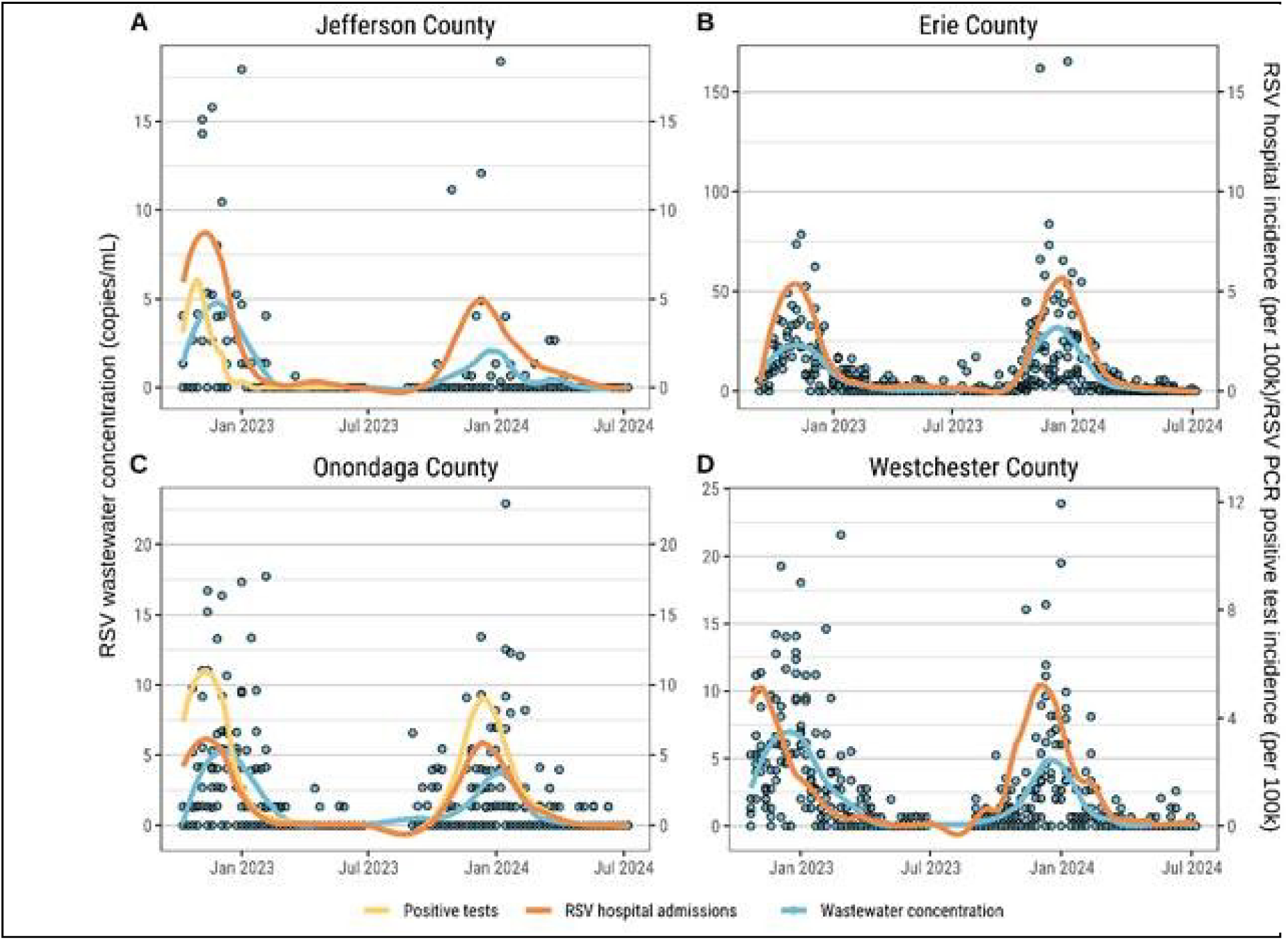
Temporal trends of RSV wastewater detection and clinical data. A) **Jefferson** County trends. NREVSS reported positive tests for RSV were only reported in the 2022 to 2023 season. B) Erie County trends. No reported RSV positive test data were available. Hospitalizations were reported for both seasons. C) Onondaga County trends. The scale for reported RSV positive incidence in Onondaga county was divided by 5 to align the scales correctly. There were roughly five times as many positive tests reported than hospital admissions in Onondaga County. Smoothed lines were calculated using locally estimated scatterplot smoothing (LOESS) with a span of 0.25. Note there are different y-axis scales for each county.

### 3.3 Accuracy and sensitivity of RSV wastewater detections

To determine the likelihood that an RSV positive detection in wastewater indicates that there are infections in the community, we compared the frequency of wastewater positive detections to the frequency of reported RSV hospitalizations. We classified each wastewater detection into four categories (true positive, false positive, true negative, and false negative) using the wastewater signal as the test. For example, true positives were times when a wastewater sample was positive in a given week that an RSV hospital admission as also reported. Using these four categories, we computed sensitivity and specificity and used these values to compute the tradeoff between the two. The tradeoff accuracy between sensitivity and specificity for an RSV positive wastewater detection to also occur during the same week as an RSV hospital admission during all weeks of testing was 84%, meaning that in this study, wastewater results had 84% probability of indicating that there would be an RSV hospitalization the same week in that community (Figure 3). We repeated this analysis for RT-PCR RSV positive clinical case data, and we found that the sensitivity of detection for an RSV wastewater detection to also occur during the same week as an RSV case report was 78% (95% CI [0.69, 0.88]) and the specificity was 89% (95% CI [0.68, 1.00]) (Table S2).

**Figure 3:**
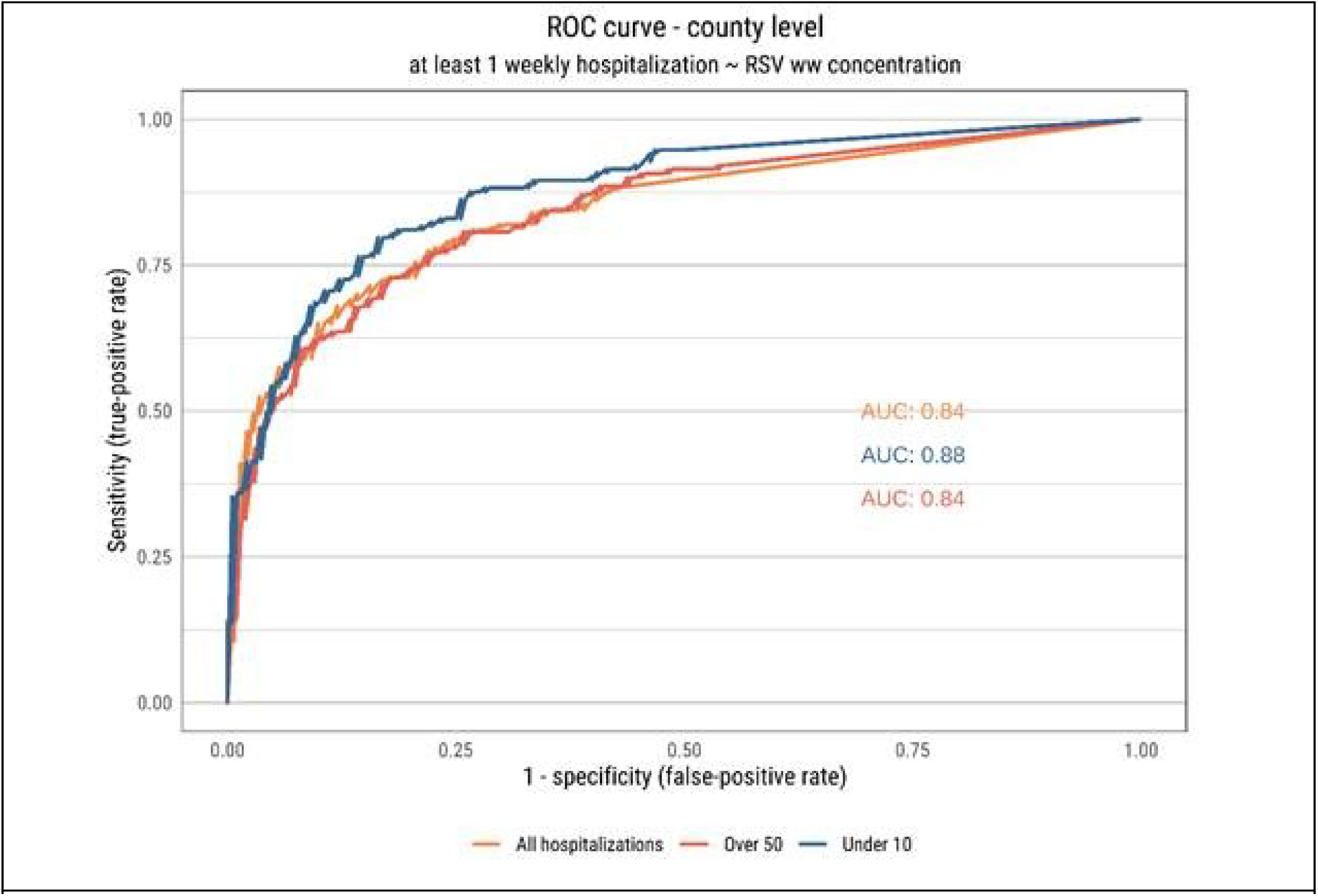
Receiver operator characteristics (ROC) curves for RSV wastewater detections. The ROC curve shows the tradeoff between sensitivity and specificity of an RSV positive detection in wastewater and the probability that the test predicted an RSV hospital admission that same week. AUC values indicate the probability that RSV wastewater detections will correctly predict RSV hospital admission of the three types that same week. Calculations were made at the county level for all hospitalizations, under 10-year-old hospital admissions and over 50-year-old hospitalizations. Sensitivity, specificity, positive predictive value, and negative predictive values for county and sewershed data are reported in Table S2.

### 3.4 Correlations between wastewater detections and clinical data

Using weekly county population-weighted averages for RSV concentration in wastewater and weekly total RSV case and hospitalization incidence, we computed Spearman correlation coefficients. Overall, correlations between RSV wastewater concentration and clinical data were strong. For cases and wastewater concentration, correlation for Jefferson County was 0.51 (95% CI [0.26, 0.64]) and, for Onondaga County, 0.72 (95% CI [0.60, 0.80]) (Table S3). Peak correlations were observed for the case data when no lead or lag time was considered; examining lead and lag between wastewater signal showed that the optimal correlation was at week zero between cases and wastewater concentration (Figure 4A). For hospital admissions and wastewater concentration, Spearman correlations were also strong ranging between 0.63 (95% CI [0.441, 0.743]) for Jefferson County and 0.85 (95% CI [0.750, 0.899]) for Erie County (Table S3). We also calculated Spearman correlations between the weekly wastewater data and weekly hospitalizations for different lag and lead times to determine if wastewater signal preceded clinical reports, or if clinical data preceded wastewater reports. When data from all ages were analyzed collectively, peak correlations were observed two weeks before wastewater levels peaked, i.e. wastewater signals lagged RSV hospital admissions (Figure 4A). In addition, for hospitalizations of children under 10 years old, wastewater data lagged by three weeks for peak correlation suggesting a longer lag for childhood incidence. However, for over 50-year-old hospitalizations, wastewater concentration led by two weeks indicating that wastewater signal preceded the rise in infections among over 50-year-olds (Figure 4A). This trend was consistent for both seasons (Figure 4B).

**Figure 4:**
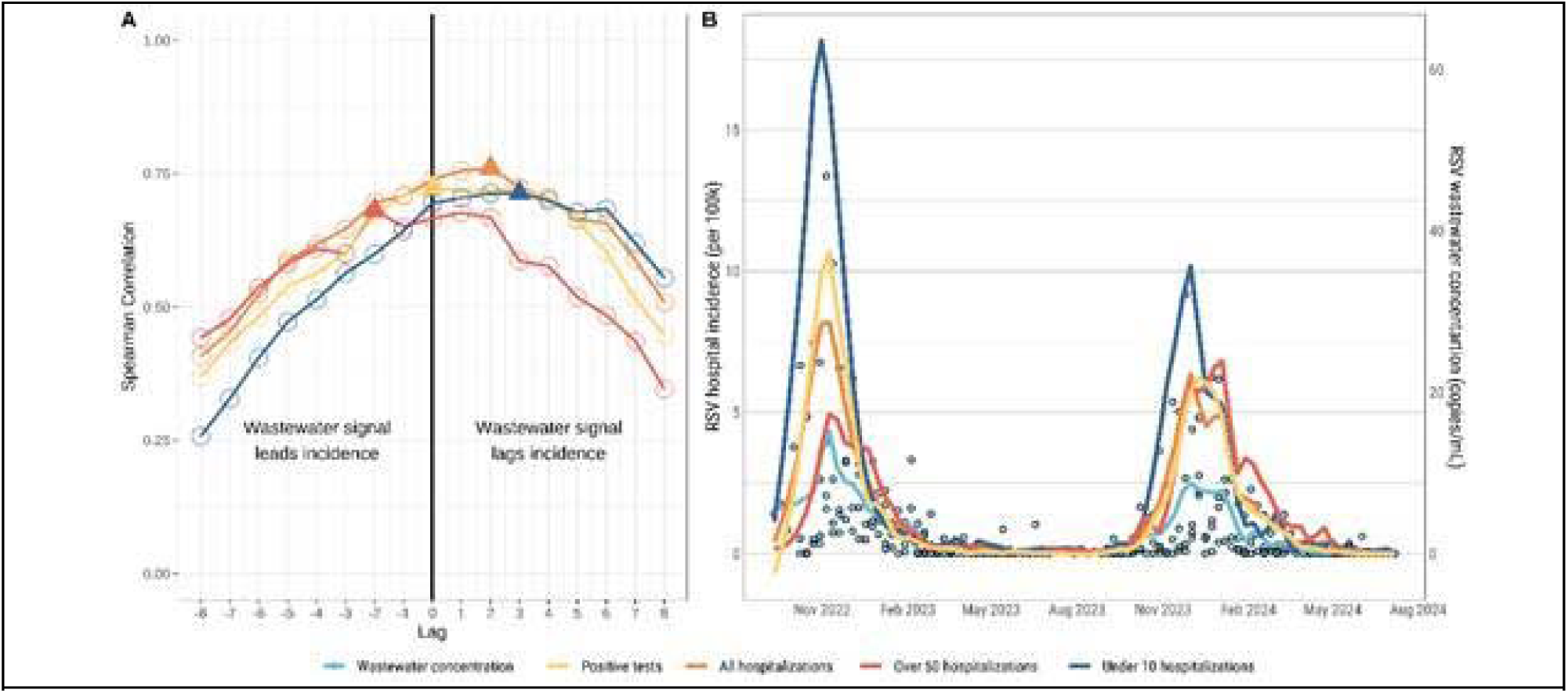
Lead time for RSV in wastewater. Wastewater values are weekly county population-weighted averages. Clinical data are weekly totals. A) Lag and lead times for wastewater concentration correlations and RSV case and hospital admissions. Triangles indicate peak correlation between lagged wastewater and case or hospitalization incidence. B) Temporal trends for RSV wastewater concentration, RSV cases, and RSV hospital admissions. Hospital admissions are all reported as incidence per 100,000 with under 10 admissions divided by three to scale the values in the plot. Case correlations and trends are for Onondaga county only and incidence of cases is divided by two to fit in the same scale as hospital admissions in panel B.

### 3.5 Wastewater as predictive of RSV transmission

Given that some lead time was observed for RSV hospitalization among over 50-year-olds, we fit predictive models to test the ability of wastewater to forecast hospitalizations among this age group. We fit a GLMM that included RSV hospitalizations in children under 10 and RSV wastewater detections as two leading indicators and we tested four models that lagged both values at similar weekly time steps (lag of zero weeks, lag of one week, and lag of two weeks). Wastewater was a significant predictor of over 50-year-old RSV hospitalizations in all wastewater only models (*P* <0.05, Table 2) and most models with both predictors (Table 2). The wastewater only model with the highest R^2^ was for a two-week lag with an R^2^ of 0.88 (β = 0.14, *P* = 0.009, Table 2).

**Table 2:** Regression results for predicting RSV hospitalizations over 50. Generalized linear mixed model results for predicting RSV hospitalizations among over 50-year-olds from wastewater and under 10 hospitalizations. Coefficients are standardized, outcome is on original scale of hospitalizations per 100k

|  |  |  |  |  |
| --- | --- | --- | --- | --- |
| **Model** | **Variable** | **Both Predictors** | **Wastewater Only** | **Hospitalizations Only** |
| No time lag | Intercept | 1.505<br>(0.3216)<br>p<0.001 | 1.512<br>(0.3624)<br>p<0.001 | 1.470 (0.3336)<br>p<0.001 |
|  | Log+1 RSV wastewater conc | 0.8062<br>(0.1494)<br>p<0.001 | 0.8759<br>(0.1493)<br>p<0.001 | - |
|  | Log+1 RSV under 10 hospitalizations | 0.3524<br>(0.1589)<br>p=0.027 | - | 0.5427 (0.1768)<br>p=0.002 |
|  | n | 337.0 | 337.0 | 337.0 |
|  | Efron R<sup>2</sup> | 0.8060 | 0.8220 | 0.8220 |
|  | AIC | 1231 | 1234 | 1257 |
| 1 week lag | Intercept | 1.506<br>(0.2633)<br>p<0.001 | 1.490<br>(0.3960)<br>p<0.001 | 1.494 (0.2672)<br>p<0.001 |
|  | Log+1 RSV wastewater conc | 0.2353<br>(0.1487)<br>p=0.114 | 0.3372<br>(0.1627)<br>p=0.038 | - |
|  | Log+1 RSV under 10 hospitalizations | 0.8808<br>(0.1607)<br>p<0.001 | - | 0.9344 (0.1640)<br>p<0.001 |
|  | n | 337.0 | 337.0 | 337.0 |
|  | Efron R<sup>2</sup> | 0.7930 | 0.8330 | 0.8050 |
|  | AIC | 1237 | 1262 | 1237 |
| 2 week lag | Intercept | 1.562<br>(0.2837)<br>p<0.001 | 1.568<br>(0.4145)<br>p<0.001 | 1.544 (0.2875)<br>p<0.001 |
|  | Log+1 RSV wastewater conc | 0.2925<br>(0.1439)<br>p=0.042 | 0.3955<br>(0.1507)<br>p=0.009 | - |
|  | Log+1 RSV under 10 hospitalizations | 0.7738<br>(0.1782)<br>p<0.001 | - | 0.8549 (0.1835)<br>p<0.001 |
|  | n | 333.0 | 333.0 | 333.0 |
|  | Efron R<sup>2</sup> | 0.7950 | 0.8450 | 0.8000 |
|  | AIC | 1220 | 1236 | 1222 |

Models with both predictors did not have as high of R^2^ values, but all were above 0.8, still suggesting high predictive power (Table 2). The highest R^2^ for a model with both predictors was at zero time lag (R^2^ = 0.84, Table 2). The highest R^2^ for a model predicted by under-10 hospitalizations only was 0.85 and also was at time lag zero (Table 2).

Table 2: Generalized linear mixed model results for predicting RSV hospitalizations among over 50-year-olds from wastewater and under 10 hospitalizations. Coefficients are standardized, outcome is on original scale of hospitalizations per 100k.

## 4 Discussion

RSV was detectable in wastewater and concentrations were highest during respiratory season which aligned with when most clinical testing and surveillance sites reported positive detections. Different quantification levels in wastewater across the counties were likely due to the two different extraction methods used with the possibility that the magnetic bead method was more sensitive, although further studies will have to corroborate this. As wastewater detections rose, RSV positive cases and reported hospitalizations also rose. Detection rates of RSV in wastewater were highest in the more urban counties of Onondaga, Erie, and Westchester, while the more rural county of Jefferson had lower detection rates. While there were the fewest RSV hospitalizations in Jefferson County, it had the highest hospitalization incidence.

General trends between wastewater concentration and clinical data matched each other well with high correlation across the two counties. There was also a high level of sensitivity and specificity in wastewater detections with regard to indicating RSV positive cases or RSV hospital admission that same week. The sensitivity levels we observed for RSV averaged around 70 to 80% and were similar to other respiratory pathogens detected in wastewater such as SARS-CoV-2, which has had reported sensitivity levels between 70% at the community level^34^ and up to 95% at the building level^35^.

RSV did not have the same early warning signal that has been observed for some other pathogens tested in wastewater like SARS-CoV-2^25,36,37^ and Hepatitis A^38^. In predictive models, RSV wastewater concentration was a strong and significant predictor of over-50-year-old RSV hospitalizations with high explanatory power up to two weeks prior to the clinical report. However, RSV wastewater concentrations were not early indicators of under-10-year-old RSV hospitalizations. This could be because infection dynamics are very different for RSV with only two subpopulations having a high risk of severe disease and prolonged infection, while most other people infected with RSV have almost no risk of severe disease and shorter infections^9^. While there have not been many studies that compare the timing of RSV infections^14^, our analysis subdividing the hospitalization data suggests that children are the first to be infected in the community and older adults are infected later in the season. The wastewater data lagging the under-10-year-old hospitalizations and leading the over-50-year-old hospitalizations reflect this dynamic. If during typical respiratory seasons, children are infected first, then the contact they have with adults in their lives leads to additional infections and shedding, which then leads to contact with older adults who become infected at higher rates toward the end of the season. Infants have been shown to shed the virus for an average of seven days with a range of one to twenty-one days, and children under one shed the most virus and have relatively low mortality at less than 0.5%^39^. Contact with children has been shown to be a key predictor of RSV infection in adults ^40^ and the average hospital stay for adults (>50 age) is three to six days with a higher mortality between 6 and 8%^41^. Thus, infections in children occur at higher incidence earlier in the season and wastewater concentration does not rise to high levels in wastewater until a few weeks later. By the time older adults are exhibiting a higher incidence, the wastewater levels are high due to an overall higher infection prevalence in the community, and this wastewater signal precedes the curve of infections among the older adult population. It is possible that children are recovering faster than adults and shedding for a shorter period of time or that infections are less severe; both of these factors have been found to cause variation in shedding times^42^.

These results show the potential for RSV monitoring in wastewater to provide early warning for burden of disease among older adults, where RSV is generally less understood and underreported^43^. While older adults experience fewer symptomatic infections, the higher mortality rate in this population means that early indicators for them could be impactful. Communities and gathering places with a high density of older populations such as churches, nursing homes, and senior centers might benefit most from the early warning. Wastewater testing also has the advantage of representing many places with on sample that might have different levels of clinical surveillance due to resource limitations^44^ and WBE data do not rely on case reports or positive diagnoses. Such early warning models have been successfully built and utilized for SARS-CoV-2^25,45^, and local health departments have identified hospitalization forecasts as some of the most valuable information derived from wastewater surveillance^46^.

While most studies that report RSV concentrations in wastewater do not report corresponding clinical data, those that do have also shown that wastewater levels lag the reported case incidence^47,48^ with one study showing mixed results where some locations have early indicator of outbreaks in wastewater and others have delayed signal^49^. Previous studies have also observed that RSV in wastewater may lag reported cases and hospitalizations but did not break out the hospitalizations into separate age groups like we do here^50^, and this could be why our study observed some early warning among sub-populations of RSV susceptible groups.

Our study has some limitations. First, many of the samples collected in the 2022 to 2023 season were stored for more than one year prior to being analyzed for RSV. While the extracted nucleic acids were stored at −80°C, there is always the possibility of degradation. Despite the time delay for PCR analysis, we were still able to detect RSV viral material in enough quantity to establish significant, positive correlations, and thus we do no think that this had a substantial effect on the study findings. Second, clinical RSV positive test data were not available for all counties in our study with only Jefferson and Onondaga counties reporting to NREVSS. Thus, our sample size for the comparison of case data with wastewater results is smaller and therefore the interpretation of results for sensitivity, specificity, PPV, and NPV for RSV case data should be done with this in mind. Findings for RSV hospitalizations are likely more accurate given that we had full coverage of both respiratory seasons and all four counties. Another limitation is that the shedding dynamics of RSV into sewer systems are poorly understood. RSV is a respiratory pathogen that does not impact the digestive tract, therefore the amount of virus shed in wastewater is typically less than for some other pathogens, such as SARS-CoV-2^51^ or polio^52^. Still, the quantity detected was more than sufficient to establish trends, predictive ability, and associations with RSV clinical data. Furthermore, the timing of shedding is likely to vary by population, though one study found that RSV can be shed for up to eleven days after infection, and up to 13% of case having prolonged shedding anywhere up to three weeks^42^.

## 5 Conclusion

Wastewater surveillance is another tool that public health agencies can use to monitor and understand transmission of RSV in their communities and can provide early warning for infections among older adults that are at high risk for hospitalization and death. RSV wastewater data are a good candidate for the creation of forecasting models due to the lead time they provide for adult hospitalizations, and the gap they can fill in surveillance. Further, clinical data often face a significant delay in reporting and wastewater data might be able to fill this gap. The data could be combined with other measures to predict hospitalizations during respiratory season.

## 6 Appendices

Supplemental tables and figures are provided in a separate document.

## 7 Credit Authorship

**Dustin T. Hill**: conceptualization, methodology, software, validation, formal analysis, investigation, data curation, writing – original draft, writing – review and editing, visualization

**Jennifer Laplante:** methodology, validation, data curation

**Sylvia Byun:** data curation; writing – review and edity

**Mohammed Alazawi:** data curation

**Donna Gowie:** data curation

**Sonali Biswas:** methodology, validation, data curation

**Latavia Hill:** methodology, validation, data curation

**Yifan Zhu:** conceptualization, writing – original draft

**Monica R. Foote:** methodology, validation

**Haley Kappus-Kron:** writing – review and editing, data curation

**Milagros Neyra Blatz:** project administration, resources, writing – review

**Ian Bradley:** methodology, writing – review and editing

**Yinyin Ye:** methodology, writing – review and editing

**Kirsten St. George**: supervision, funding acquisition, conceptualization, project administration

**David A. Larsen:** supervision, funding acquisition, conceptualization, project administration, writing – review and editing

## 9 Acknowledgements

We would like to thank the extraction laboratories for processing samples including Quadrant Biosciences, and University at Buffalo. We would also like to thank the New York State Wastewater Surveillance Network for coordinating sample collection and shipping. Also, thank you to Bryon Backenson and Bridget Anderson from the New York State Bureau of Communicable Disease Control. We also are grateful to the help of operators at wastewater treatment sites that contributed samples to the program.

## 10 Data availability statement

County aggregated wastewater concentrations, RSV cases, and RSV hospital admissions are available on the GitHub page associated with this paper. All code used to produce the figures for the analysis are also included. https://github.com/nys-wwsn/rsv-wastewater-pilot

## 11 Disclaimer

The content of this publication is solely the responsibility of the authors and does not necessarily represent the official views of the CDC, HRI, or the NYS Department of Health. The authors have no conflicts of interest to disclose

## 12 Declarations

Generative AI was not used for writing or reviewing this manuscript. The generative AI software Claude was used to assist in pulling age-adjusted population totals from the tidycensus R package and in cleaning R code for creating tables in a publishable format. Generative AI was not used for data analysis.

