## Supplementary material for "RSV concentration in wastewater and trends among incident hospitalizations in children and older adults"

**Supplemental materials**

**Supplemental text**

The main text reports the results for all weeks of the year because there was not much difference in RSV detection in wastewater during all weeks as opposed to only during respiratory season (weeks 40 to 20). Looking at respiratory season only, sensitivity for a wastewater detection to correspond to a hospitalization that same week at the sewershed level was 0.72 (95% CI [0.68, 0.75]). During all weeks, sensitivity for a wastewater detection to correspond to a hospitalization that same week at the sewershed level was 0.68 (95% CI [0.64,0.71]). There was similarly little difference at the county level. During respiratory season, sensitivity for a wastewater detection to correspond to a hospitalization that same week at the county level was 0.85 (95% CI [0.80,0.90]). During all weeks, sensitivity for a wastewater detection to correspond to a hospitalization that same week at the county level was 0.82 (95% CI [0.77,0.86]). Values for sensitivity, specificity, PPV, and NPV during all weeks of the year are reported in Table S1. Values for respiratory season only are reported in Table S6 and Figure S1.

**Tables**

Table S1: Descriptive statistics for RSV detection rates split between summer months (Weeks 21-39), respiratory season (Weeks 40 to 20), all weeks of the year.

| **season** | **county** | **N sewersheds** | **N samples** | **mean** | **min** | **median** | **max** |
| --- | --- | --- | --- | --- | --- | --- | --- |
| Weeks 21 to 39 | Erie | 7 | 276 | 13.05 | 3.57 | 11.76 | 22.73 |
| Weeks 21 to 39 | Jefferson | 3 | 59 | 0.00 | 0.00 | 0.00 | 0.00 |
| Weeks 21 to 39 | Onondaga | 7 | 118 | 8.46 | 0.00 | 11.76 | 11.76 |
| Weeks 21 to 39 | Westchester | 7 | 223 | 14.78 | 9.68 | 15.38 | 21.21 |
| Weeks 40 to 20 | Erie | 7 | 504 | 56.58 | 43.14 | 50.00 | 77.27 |
| Weeks 40 to 20 | Jefferson | 3 | 230 | 22.37 | 20.34 | 21.31 | 25.45 |
| Weeks 40 to 20 | Onondaga | 7 | 417 | 36.53 | 18.18 | 39.06 | 51.56 |
| Weeks 40 to 20 | Westchester | 7 | 835 | 44.37 | 38.46 | 43.80 | 52.42 |
| All weeks | Erie | 7 | 780 | 41.12 | 29.21 | 36.51 | 59.09 |
| All weeks | Jefferson | 3 | 289 | 17.80 | 15.79 | 17.33 | 20.29 |
| All weeks | Onondaga | 7 | 535 | 30.44 | 14.29 | 33.33 | 43.21 |
| All weeks | Westchester | 7 | 1,058 | 38.14 | 33.78 | 36.77 | 43.95 |

Table S2 Sensitivity, specificity, PPV, and NPV of RSV wastewater surveillance

| **aggregation** | **outcome** | **group** | **value** | **SE** | **95% LL** | **95% UL** |
| --- | --- | --- | --- | --- | --- | --- |
| sewershed | All hospitalizations | sensitivity | 0.68 | 0.02 | 0.64 | 0.71 |
| sewershed | All hospitalizations | specificity | 0.72 | 0.01 | 0.70 | 0.75 |
| sewershed | All hospitalizations | ppv | 0.56 | 0.02 | 0.53 | 0.60 |
| sewershed | All hospitalizations | npv | 0.81 | 0.01 | 0.79 | 0.83 |
| sewershed | Under 10 | sensitivity | 0.71 | 0.02 | 0.67 | 0.76 |
| sewershed | Under 10 | specificity | 0.68 | 0.01 | 0.65 | 0.70 |
| sewershed | Under 10 | ppv | 0.40 | 0.02 | 0.36 | 0.43 |
| sewershed | Under 10 | npv | 0.89 | 0.01 | 0.87 | 0.91 |
| sewershed | Over 50 | sensitivity | 0.71 | 0.02 | 0.66 | 0.75 |
| sewershed | Over 50 | specificity | 0.67 | 0.01 | 0.64 | 0.69 |
| sewershed | Over 50 | ppv | 0.37 | 0.02 | 0.34 | 0.41 |
| sewershed | Over 50 | npv | 0.89 | 0.01 | 0.87 | 0.91 |
| county | cases | sensitivity | 0.78 | 0.05 | 0.69 | 0.88 |
| county | cases | specificity | 0.89 | 0.10 | 0.68 | 1.09 |
| county | cases | ppv | 0.98 | 0.02 | 0.95 | 1.02 |
| county | cases | npv | 0.33 | 0.10 | 0.14 | 0.52 |
| county | All hospitalizations | sensitivity | 0.82 | 0.02 | 0.77 | 0.86 |
| county | All hospitalizations | specificity | 0.66 | 0.05 | 0.56 | 0.76 |
| county | All hospitalizations | ppv | 0.88 | 0.02 | 0.84 | 0.92 |
| county | All hospitalizations | npv | 0.54 | 0.05 | 0.44 | 0.64 |
| county | Under 10 | sensitivity | 0.87 | 0.02 | 0.82 | 0.91 |
| county | Under 10 | specificity | 0.54 | 0.04 | 0.46 | 0.62 |
| county | Under 10 | ppv | 0.73 | 0.03 | 0.68 | 0.79 |
| county | Under 10 | npv | 0.74 | 0.04 | 0.65 | 0.82 |
| county | Over 50 | sensitivity | 0.87 | 0.02 | 0.82 | 0.91 |
| county | Over 50 | specificity | 0.56 | 0.04 | 0.47 | 0.64 |
| county | Over 50 | ppv | 0.75 | 0.03 | 0.70 | 0.81 |
| county | Over 50 | npv | 0.73 | 0.04 | 0.64 | 0.81 |

Table S3: Spearman correlations for each county for weekly RSV wastewater detections and weekly RSV PCR positive tests (cases) or weekly RSV hospitalizations.

| **Aggregation** | **values tested** | **Spearman's r** | **95% LL** | **95% UL** | **Observations (n)** |
| --- | --- | --- | --- | --- | --- |
| Jefferson | log+1(Cases ~ concentration) | 0.507 | 0.263 | 0.642 | 83 |
| Onondaga | log+1(Cases ~ concentration) | 0.723 | 0.595 | 0.8 | 82 |
| Erie | log+1(Hospitalizations ~ concentration) | 0.847 | 0.75 | 0.899 | 94 |
| Jefferson | log+1(Hospitalizations ~ concentration) | 0.628 | 0.441 | 0.743 | 83 |
| Onondaga | log+1(Hospitalizations ~ concentration) | 0.655 | 0.487 | 0.748 | 82 |
| Westchester | log+1(Hospitalizations ~ concentration) | 0.739 | 0.637 | 0.807 | 82 |

**Table S4:** ICD 10 codes used in the analysis

| **ICD 10 code type** | **Coding System** | **Codes*** | **Description** |
| --- | --- | --- | --- |
| **All diagnoses** | ICD-10 | J12.1 | RSV Pneumonia |
| **All diagnoses** | ICD-10 | J21.0 | RSV Acute Bronchiolitis |
| **All diagnoses** | ICD-10 | J21.5 | RSV Bronchitis |
| **All diagnoses** | ICD-10 | B97.4 | RSV as cause of other diseases |

Table S5 Sensitivity, specificity, PPV, and NPV of RSV wastewater surveillance during respiratory season (Weeks 40-20)

| **season_name** | **aggregation** | **outcome** | **group** | **value** | **SE** | **95% LL** | **95% UL** |
| --- | --- | --- | --- | --- | --- | --- | --- |
| Weeks 40 to 20 | sewershed | All hospitalizations | sensitivity | 0.72 | 0.02 | 0.68 | 0.75 |
| Weeks 40 to 20 | sewershed | All hospitalizations | specificity | 0.66 | 0.02 | 0.62 | 0.69 |
| Weeks 40 to 20 | sewershed | All hospitalizations | ppv | 0.60 | 0.02 | 0.56 | 0.63 |
| Weeks 40 to 20 | sewershed | All hospitalizations | npv | 0.76 | 0.02 | 0.73 | 0.79 |
| Weeks 40 to 20 | sewershed | Under 10 | sensitivity | 0.75 | 0.02 | 0.71 | 0.79 |
| Weeks 40 to 20 | sewershed | Under 10 | specificity | 0.60 | 0.02 | 0.57 | 0.63 |
| Weeks 40 to 20 | sewershed | Under 10 | ppv | 0.42 | 0.02 | 0.39 | 0.46 |
| Weeks 40 to 20 | sewershed | Under 10 | npv | 0.86 | 0.01 | 0.83 | 0.89 |
| Weeks 40 to 20 | sewershed | Over 50 | sensitivity | 0.72 | 0.02 | 0.68 | 0.77 |
| Weeks 40 to 20 | sewershed | Over 50 | specificity | 0.58 | 0.02 | 0.55 | 0.61 |
| Weeks 40 to 20 | sewershed | Over 50 | ppv | 0.40 | 0.02 | 0.36 | 0.43 |
| Weeks 40 to 20 | sewershed | Over 50 | npv | 0.85 | 0.01 | 0.82 | 0.88 |
| Weeks 40 to 20 | county | cases | sensitivity | 0.80 | 0.05 | 0.70 | 0.90 |
| Weeks 40 to 20 | county | cases | specificity | 1.00 | 0.00 | 1.00 | 1.00 |
| Weeks 40 to 20 | county | cases | ppv | 1.00 | 0.00 | 1.00 | 1.00 |
| Weeks 40 to 20 | county | cases | npv | 0.07 | 0.07 | -0.06 | 0.21 |
| Weeks 40 to 20 | county | All hospitalizations | sensitivity | 0.85 | 0.02 | 0.80 | 0.90 |
| Weeks 40 to 20 | county | All hospitalizations | specificity | 0.62 | 0.08 | 0.47 | 0.78 |
| Weeks 40 to 20 | county | All hospitalizations | ppv | 0.93 | 0.02 | 0.90 | 0.97 |
| Weeks 40 to 20 | county | All hospitalizations | npv | 0.41 | 0.07 | 0.28 | 0.54 |
| Weeks 40 to 20 | county | Under 10 | sensitivity | 0.90 | 0.02 | 0.85 | 0.94 |
| Weeks 40 to 20 | county | Under 10 | specificity | 0.46 | 0.06 | 0.36 | 0.57 |
| Weeks 40 to 20 | county | Under 10 | ppv | 0.78 | 0.03 | 0.73 | 0.84 |
| Weeks 40 to 20 | county | Under 10 | npv | 0.68 | 0.06 | 0.56 | 0.80 |
| Weeks 40 to 20 | county | Over 50 | sensitivity | 0.87 | 0.02 | 0.82 | 0.92 |
| Weeks 40 to 20 | county | Over 50 | specificity | 0.46 | 0.06 | 0.34 | 0.57 |
| Weeks 40 to 20 | county | Over 50 | ppv | 0.82 | 0.03 | 0.76 | 0.87 |
| Weeks 40 to 20 | county | Over 50 | npv | 0.55 | 0.07 | 0.42 | 0.68 |

**Figures**

**Figure S1:** Sensitivity, specificity, PPV, NPV values for counties and sewersheds for all comparisons.

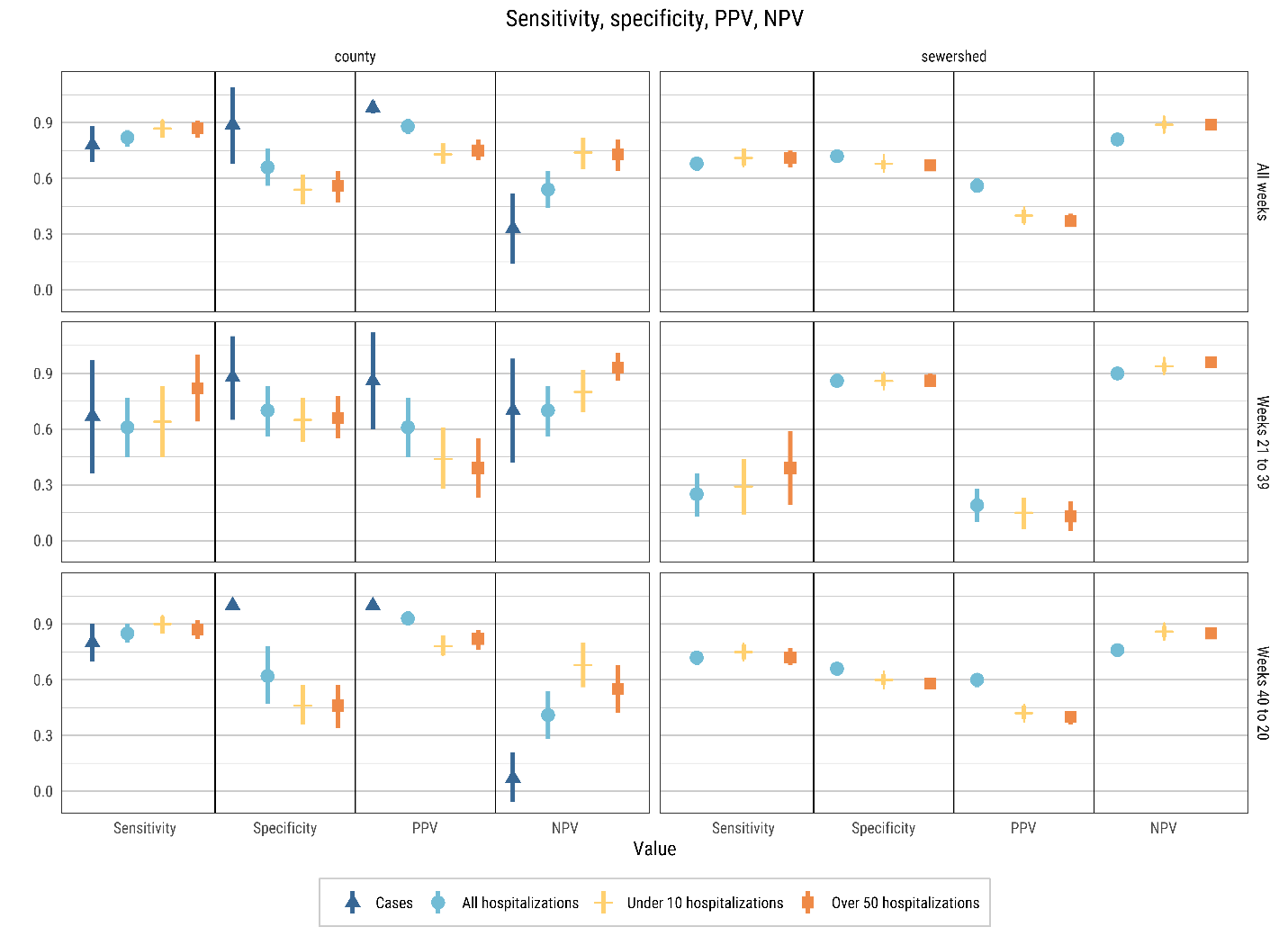
